# Self-reported sleep duration and timing: A methodological review of event definitions, context, and timeframe of related questions

**DOI:** 10.1101/2020.09.09.20191379

**Authors:** Rebecca Robbins, Stuart F. Quan, Laura K. Barger, Charles A. Czeisler, Maya Frazy-Witzer, Matthew D. Weaver, Ying Zhang, Susan Redline, Elizabeth B. Klerman

## Abstract

**Background:** Clinical practice guidelines and population health recommendations are derived from studies that include self-reported data. Small semantic differences in question wording and response scales, may significantly affect the response. We conducted a methodological review to assess the variation in event definition(s), context (i.e., work-versus free-day), and timeframe of sleep timing and duration questions.

**Methods:** We queried multiple databases of sleep, medicine, public health, and psychology for survey-based studies and/or publications with sleep duration and/or timing questions. The text of these questions was extracted and thematically analyzed by two trained coders.

**Results:** We identified 49 surveys that included sleep duration and/or timing questions. Sample sizes of participants in the reviewed publications using these surveys ranged from 93 to 1,185,106. For sleep duration questions, participants were asked to report nocturnal sleep (22/43), sleep in the past 24-hours (15/43), their major sleep episode (3/43), or no event definition was given (3/43). Among bedtime questions, participants were asked to report the time into bed (17/40), fall asleep time (10/40), or first attempt to sleep (13/40). For wake time questions, participants were asked to report their out of bed time (3/36), the time they get up (6/36), or their wake up time (27/36). Context (e.g., work versus free day) guidance was provided in 18/43 major sleep duration questions, 31/40 bedtime questions, and 30/36 wake time questions. Timeframe (e.g., the past 4 weeks) was provided in 6/43 major sleep episode duration questions, 9/40 bedtime questions, and 3/36 wake time questions. Among the surveys analyzed, only one question asked about the method of awakening (e.g., spontaneously or by alarm clock), 15 questions assessed sleep latency, and eight measured napping.

**Conclusion:** There is large variability in the event definition(s), context, and timeframe of questions relating to duration, bed/wake times, latency, and napping. This work may inform future efforts at data harmonization for meta-analyses, provide options for question wording for researchers and clinicians, and be used to identify candidate questions for future surveys.

## Introduction

Researchers and clinicians across disciplines have become aware of the importance of sleep timing and duration on multiple health and safety outcomes.^1–3^ Public health recommendations for sleep duration and timing usually rely partially on self-reported data,^4,5^ Although it is clear that differences in the specific wording used to collect information about sleep timing and duration would be expected to affect responses, high variation in sleep duration and timing survey questions has persisted in sleep research.

Survey designers must carefully balance several factors as they develop or select survey questions. First, designers must consider *event definition(s)*. For example, bedtime may be assessed by asking a participant to report when they “go to bed” or, alternatively, when they “fall asleep.” If a participant goes to bed but then reads or watches television before initiating sleep, asking this individual when they “go to bed” and using this as the marker of sleep onset will overestimate sleep duration. On the other hand, individuals may have difficulty knowing the precise time they fall asleep, and their perceived sleep quality might bias such responses. Second, designers must consider *context*. Sleep timing and duration have been shown to vary significantly between work and free days,^6,7^ so it is optimal to capture context with separate questions where possible. Third, survey designers must consider *timeframe*. Whereas some surveys may ask participants to report sleep duration and/or timing on a “typical night,” others ask for sleep duration and/or timing in a specified timeframe (e.g., “in the past 4 weeks”). Responses may not be comparable between study participants who may consider different reference periods as they make their response. Finally, the survey designer must balance the detail/depth of information sought with other survey constraints (e.g., the number of questions asked may influence survey completion rate). Therefore, considering the potential effects of variations in event definition(s), timeframe, and context of questions is critical in using responses to queries on sleep duration and timing.

Goals of this work were to improve awareness of the variation in event definition(s), context, and timeframe across surveys that collect information on sleep duration and timing, and to highlight options for survey designers to consider. This work should also support future efforts to standardize questions pertaining to sleep duration and timing and thus enable meta-analyses across studies to improve understanding of sleep’s role in health and safety.

## Methods

We conducted a methodological review of questions measuring sleep duration and timing in adults. In our review, we identified databases and repositories managed by organizations in sleep, medicine, public health, and psychology. We downloaded and then qualitatively analyzed and categorized the relevant questions, their wording, and response scales.

Studies were also classified by whether their aim included validation. A study that aimed to validate questions was one that examined psychometric properties of the questions with another objective measurement, such as wrist-worn actigraphy. A non-validated survey administered questions without the validation efforts of such a study.

### Identifying relevant questions

We identified six databases or repositories that include sleep, medicine, public health, and psychology disciplines, and were managed by academic or professional institutions (details and links in Appendix): the Sleep Research Society (SRS); the National Sleep Research Resource (NSRR); the Institute for Social Research at the University of Michigan (ICSPR); the American Psychological Association Health and Psychological Instruments (HAPI); the American Thoracic Society (ATS); and the National Institutes of Health (NIH).

For each database or repository, we searched for studies with questions measuring sleep duration and timing. ICSPR, ATS, and HAPI had search tools that we used to identify relevant studies; we used the following search term to identify relevant studies: “sleep,” “sleep duration,” “sleep timing,” “bedtime,” “wake time,” “bed,” “wake,” “woke up,” “alarm clock,” “asleep,” and “nap.” For databases without a search tool, each study in the database was individually reviewed for relevant sleep-related questions.

We noted which studies were conducted with the primary aim of validating a survey using steps to ascertain the psychometric properties of the questions with another objective measurement versus those studies that administered survey questions to assess sleep duration and/or timing without the primary aim of validating the survey questions themselves.

The studies identified using the above procedures were cross-referenced in two additional steps to ensure our list was comprehensive. First, we reviewed a textbook with more than 100 sleep surveys for sleep duration and timing questions.^8^ Second, an expert team (EBK, CAC, SFQ, SR, LKB) reviewed the list of studies that emerged from all the above steps and identified additional missing studies that were added to our review. Finally, we removed surveys that targeted children and adolescents or were not in English.

### Coding

The exact text of questions assessing sleep duration and timing from each study were extracted and reviewed. Two independent coders (RR and MFW) qualitatively analyzed the questions to identify domains used to measure sleep duration and timing. The coders removed questions that measured domains that did not specifically pertain to the duration or timing of sleep (e.g., about sleep quality or sleep difficulty); questions asking individuals for their preferred amount of sleep as opposed to their actual sleep; and questions that asked participants to report the number of hours of sleep after which they are able to wake up and feel refreshed. The broad sleep duration and timing domains used included: sleep duration; bedtimes; wake times and/or method of waking; sleep latency; and naps.

For sleep duration, we were interested in questions assessing total sleep across the 24-hour day, nocturnal sleep, or the major sleep episode (i.e., the longest period of time after falling asleep until the time sleep ends and the person chooses to remain awake); questions assessing naps or periods of sleep that were less than four hours; and questions assessing sleep latency (i.e., the time it takes to fall asleep). For sleep timing, we were interested in questions assessing bedtimes and wake times, and sleep latency. Question text of “what time do you go to bed” and “what time do you get out of bed,” was classified as time in bed while text of “what time did you fall asleep” and “what time did you wake up” was classified as the major sleep episode (Figure 1).^9^

**Figure 1.**
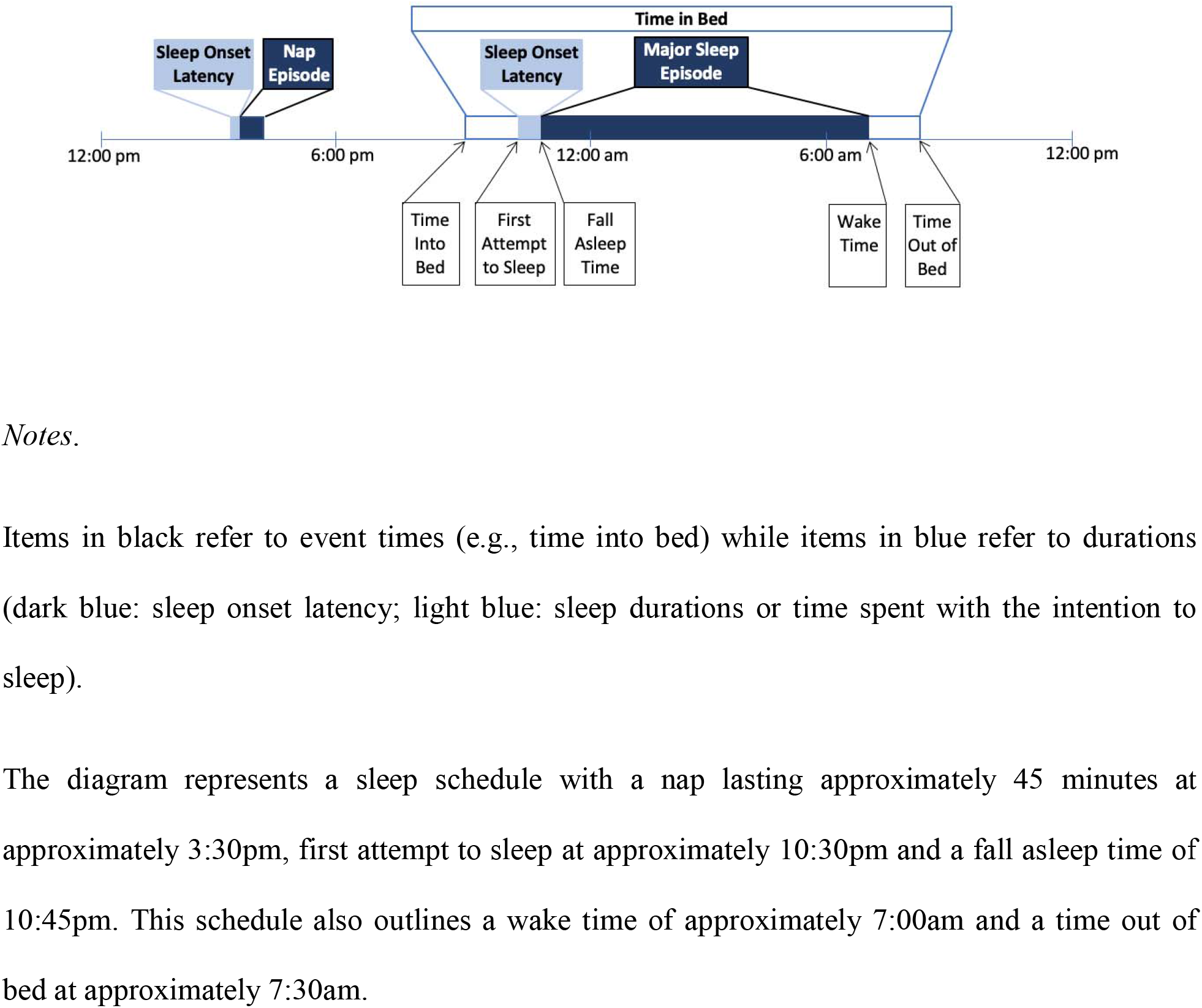
A graphical display of sleep duration and timing terminology used in this report. *Notes*. Items in black refer to event times (e.g., time into bed) while items in blue refer to durations (dark blue: sleep onset latency; light blue: sleep durations or time spent with the intention to sleep). The diagram represents a sleep schedule with a nap lasting approximately 45 minutes at approximately 3:30pm, first attempt to sleep at approximately 10:30pm and a fall asleep time ofof10:45pm. This schedule also outlines a wake time of approximately 7:00am and a time out of bed at approximately 7:30am.

Coders first separated all the questions into the above-detailed sleep domains, then applied appropriate codes to summarize the approaches (e.g., event definition, context, timeframe) taken in each question. For event definitions, coders identified content present in the questions that directed the participant and were supplemental to the clear, primary purpose of the question. For instance, a question such as “How many hours of sleep do you get?” would not be coded as featuring additional event definition(s), whereas a question such as “How many hours of sleep do you get at night (<u>this may be different than the time you spend in bed</u>)?” would be coded as featuring event definition detail to subtract time in bed not sleeping. For context, coders identified questions that asked participants to distinguish between work- and free-day sleep duration and sleep timing-related responses. Questions that provided any context cues would be categorized as distinguishing between work and free day sleep duration and/or timing. For instance, some surveys only ask one question about sleep duration on weekdays, which would be categorized as representing context cues. Finally, coders identified questions that provided participants with guidance on the timeframe they should report on their sleep. For instance, the question “<u>In the past 4 weeks</u>, how many hours have you slept at night?” provides participants with timeframe input to guide their sleep duration and/or timing response.

The coders met regularly to review and discuss their coding, agree upon a shared codebook, and then returned to coding until all disagreements in codes were rectified and all questions had been coded. The exact wording of all questions coded is included in Tables 2-5.

## Results

Our search yielded 49 studies assessing sleep duration and/or timing (9 validated surveys and 40 non-validated surveys). Sample sizes of the studies ranged from 93 to 1,185,10610. Approximately one third of studies in our sample (39%) measured only one of the five identified domains, 18% measured 2 domains, 29% measured 3 domains, 12% measured 4 domains, and 2% of studies measured all five domains (Table 1).

**Table 1.**
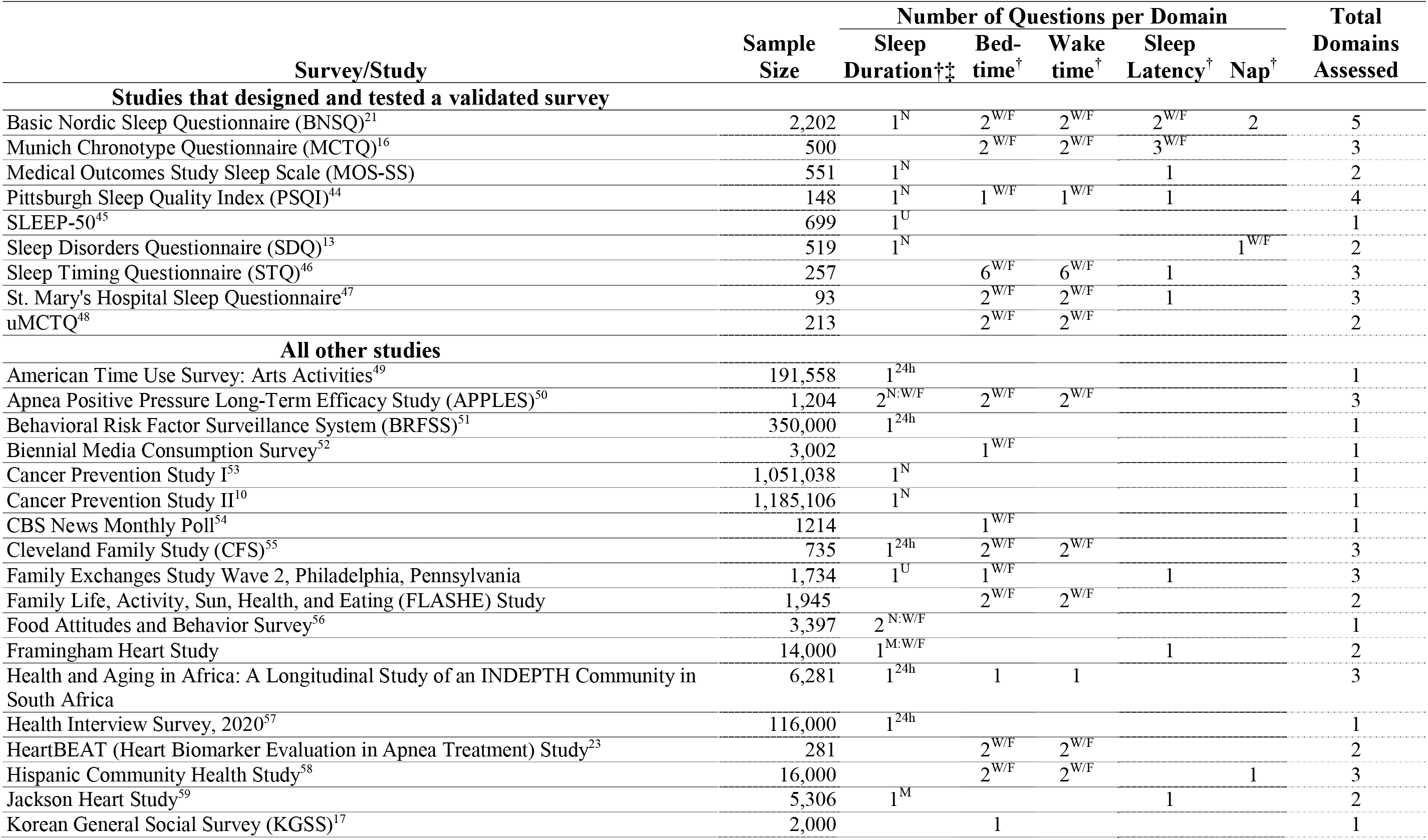
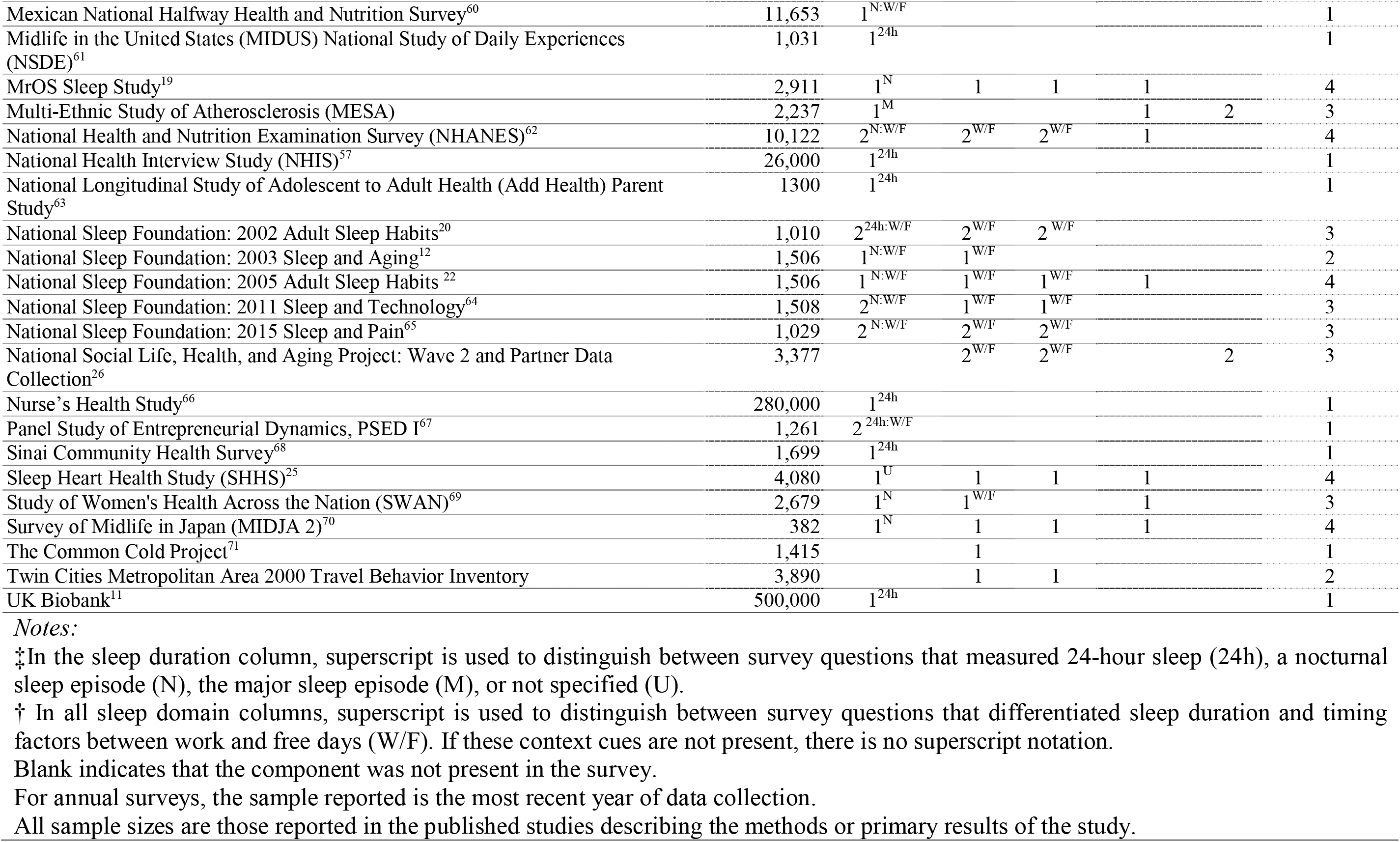
Summary characteristics of the sleep timing and duration domains assessed.

### Sleep Duration (Table 2)

We distinguish between studies that measured nocturnal sleep (e.g., “How many hours do you usually sleep per night?”), 24-hour sleep duration (e.g., “On average, how many hours of sleep do you get in a 24-hour period?”) and major sleep episode (e.g., “How much sleep do you usually get at night (or your main sleep period)?”). Among the 43 questions measuring sleep duration, 22 questions asked participants to report nocturnal sleep, 15 questions asked participants to report their sleep in the past 24 hours, three questions asked participants to report their major sleep episode, while three questions were unclear or unspecified (e.g., “I sleep __ hours”).

**Table 2.**
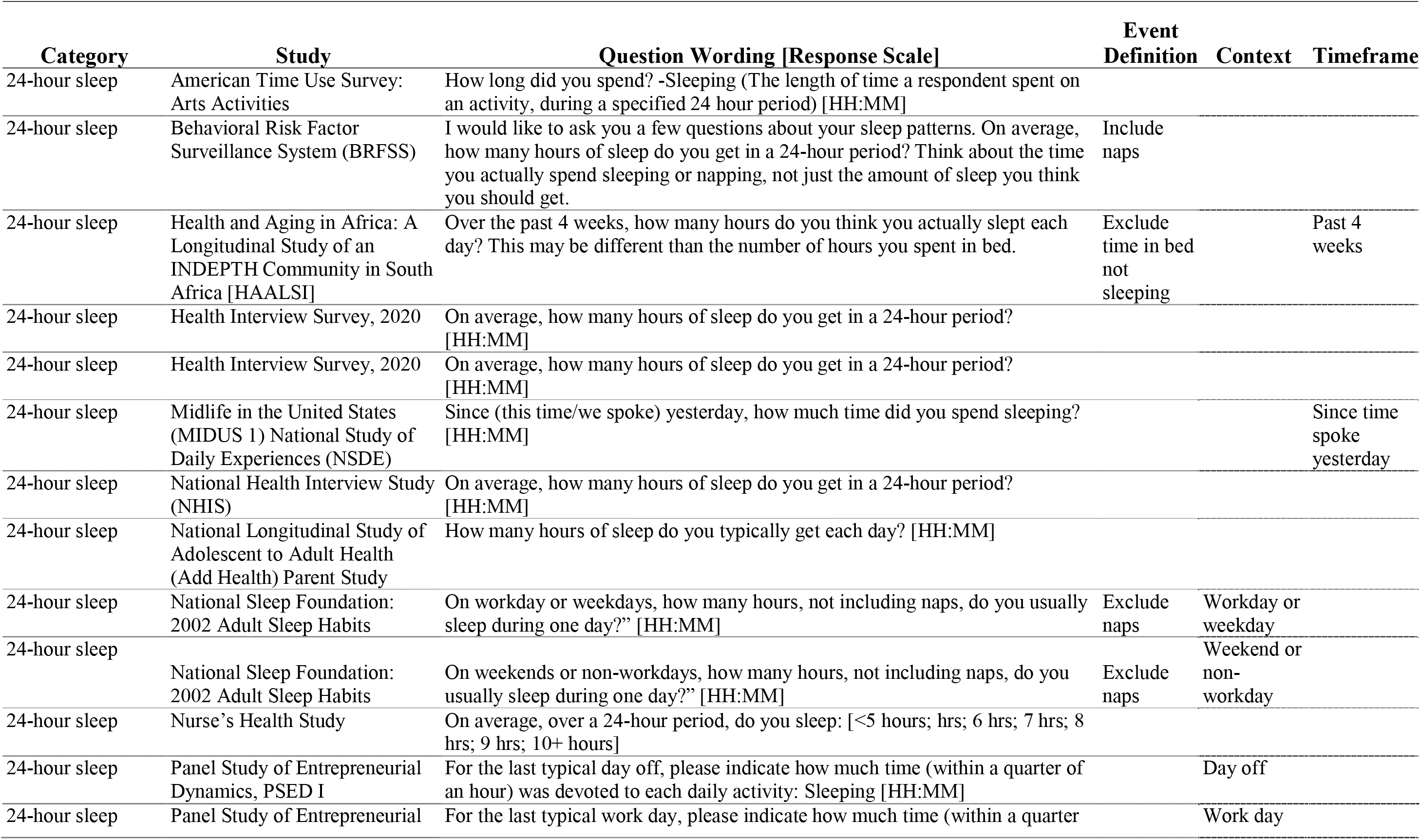
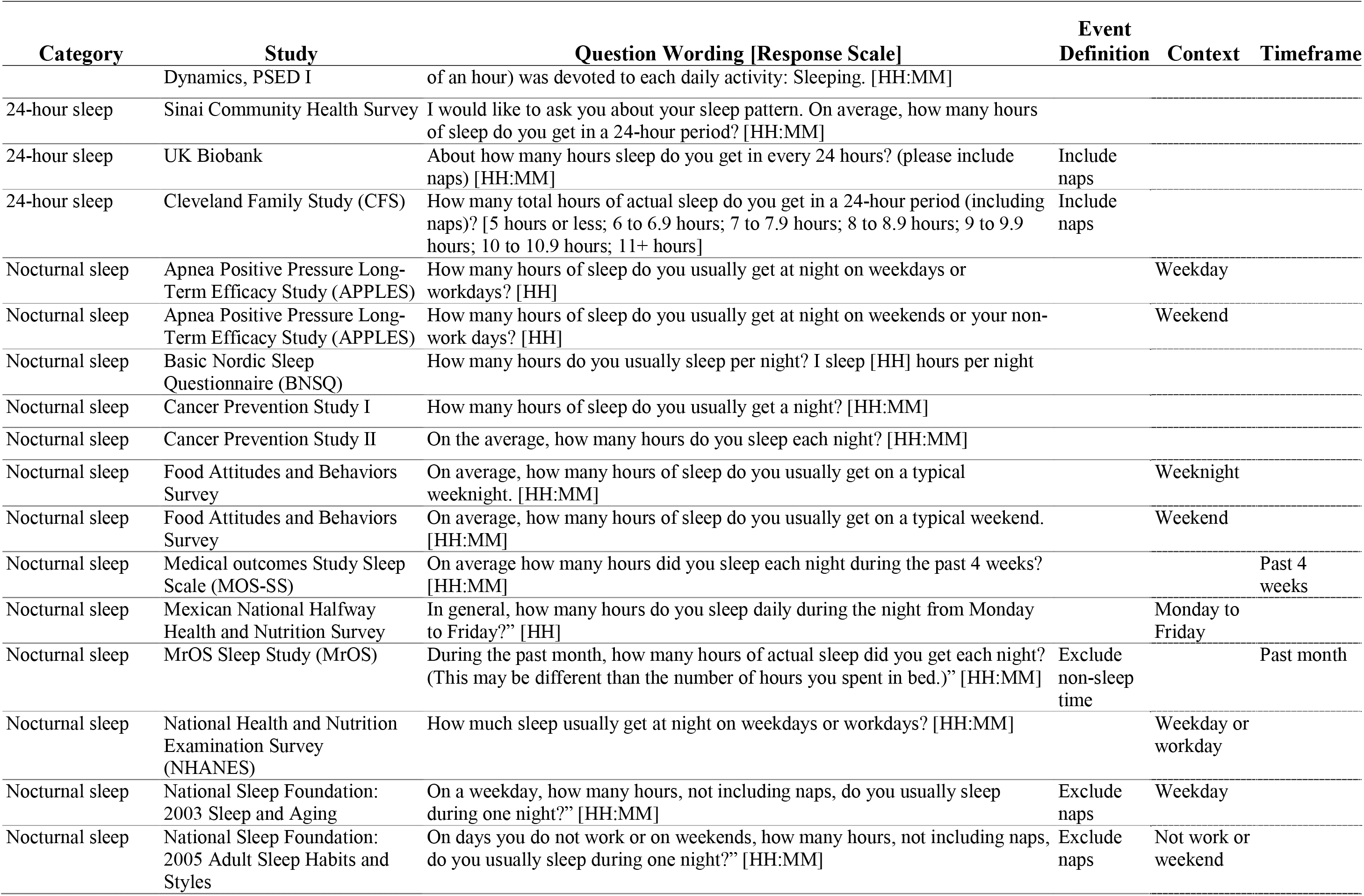
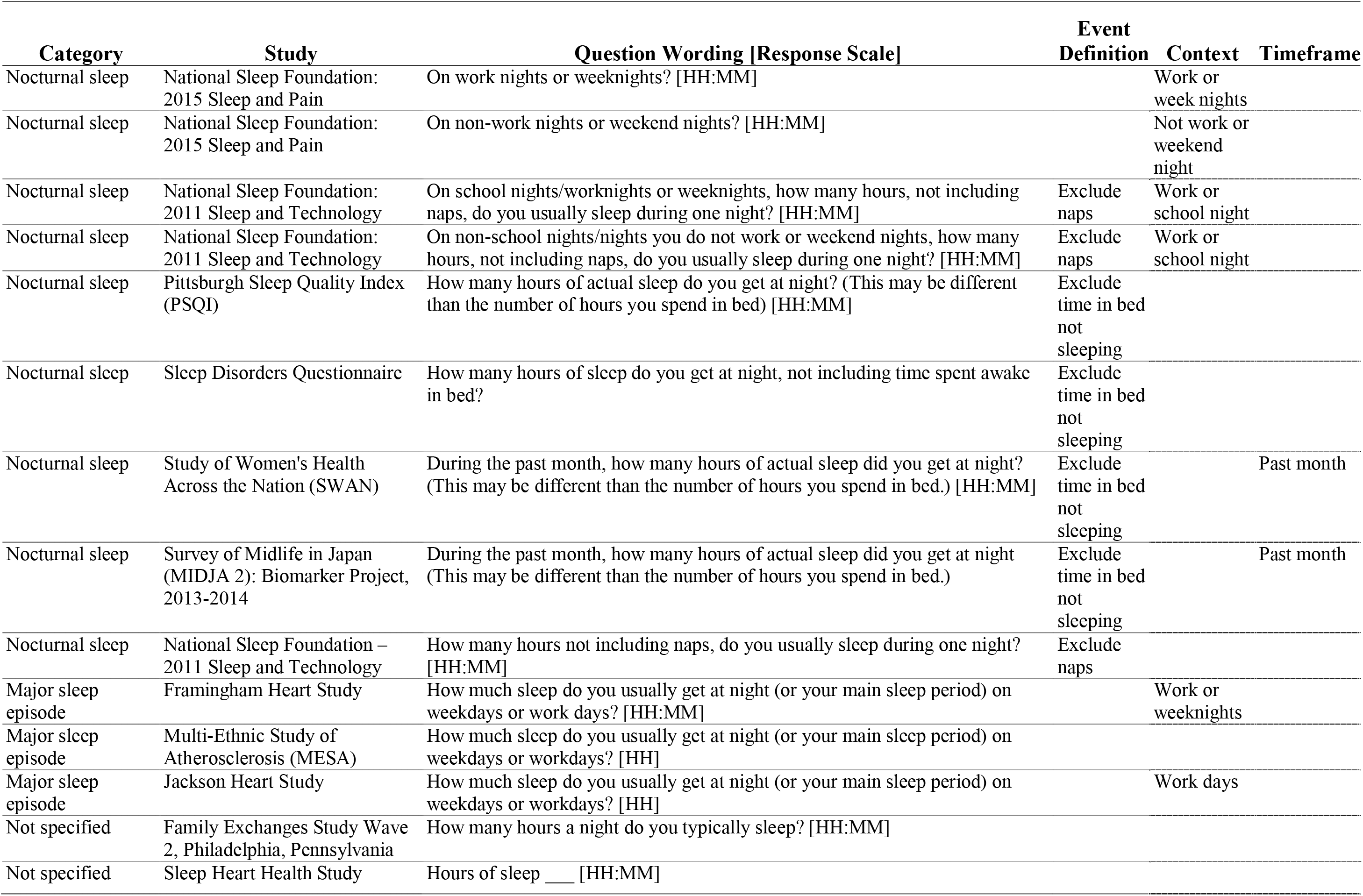
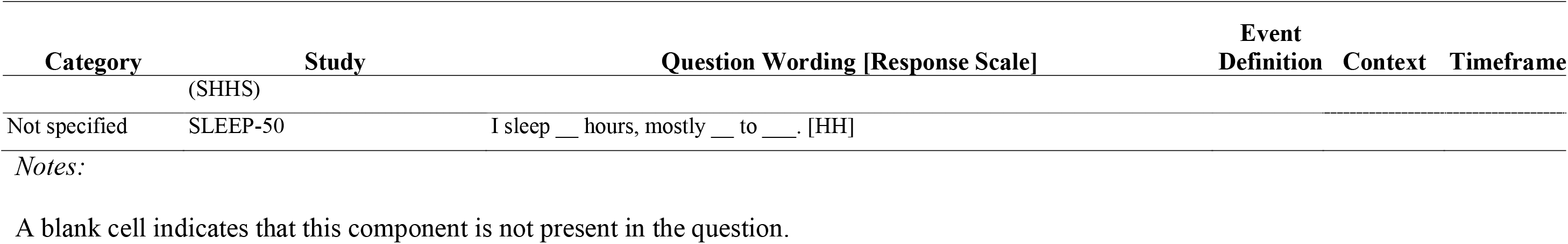
Sleep duration questions.

Sixteen duration questions provided respondents with event definition details. Specifically, 3 questions asked participants to include naps in their sleep duration responses (e.g., United Kingdom, UK, Biobank: “About how many hours sleep do you get in every 24 hours? Please include naps”),^11^ while 7 questions asked participants not to include naps (e.g., National Sleep Foundation 2003 Sleep and Aging Survey: “On a weekday, how many hours, not including naps, do you usually sleep during one night?”),^12^ and 6 surveys asked participants to subtract time spent in bed not sleeping from sleep duration calculations (e.g., the Sleep Disorders Questionnaire: “How many hours of sleep do you get at night, not including time spent awake in bed?”).^13^

Among the sleep duration questions, 18 asked participants to distinguish between work-and free-day sleep (e.g., Framingham Heart Study: How much sleep do you usually get at night (or your main sleep period) on weekdays or work days?”)^14^ and 6 questions asked participants to report their sleep in a specific timeframe (e.g., the Medical outcomes Study Sleep Scale: “On average how many hours did you sleep each night during the past 4 weeks?”).^15^

### Bedtime and Wake Time and/or Method of Waking (Table 3)

We identified 40 distinct questions assessing bedtime. Seventeen questions asked participants to report “time got into bed” (e.g., Munich Chronotype Questionnaire: “On nights before free days, I go to bed at [ ] o clock …?”),^16^ 10 questions asked participants their fall asleep time (e.g., Korea General Social Survey: “At about what time did you go to sleep yesterday?”),^17^ and 13 questions asked participants when they first attempt to fall asleep (e.g., Family Life, Activity, Sun, Health, and Eating (FLASHE) Study: “What time do you usually go to bed in the evening (turn out the lights in order to go to sleep)?”).^18^ Nine surveys asked participants to report their usual bedtimes in a specified timeframe (e.g., the MrOS Sleep Study: “During the past month, what time have you usually gone to bed at night?”).^19^ Twenty-five questions asked participants to distinguish between work- and free-day bedtimes (e.g., National Sleep Foundation 2002 Adult Sleep Habits: “At what time do you usually go to bed on nights before workdays or weekdays?”).^20^

**Table 3.** Sleep timing questions.

| **Category** | **Study** | **Question Wording [Response Scale]** | **Event Definition** | **Context** | **Timeframe** |
| --- | --- | --- | --- | --- | --- |
| Bedtime | Cleveland Family Study | During the PAST MONTH, what TIME (on average) have you gone to bed on weekdays (first closed eyes in attempt to fall asleep)” [HH:MM am/pm] | Attempt to sleep | Weekday | Past month |
| Bedtime | Cleveland Family Study | During the PAST MONTH, what TIME (on average) have you gone to bed on weekends (first closed eyes in attempt to fall asleep)” [HH:MM am/pm] | Attempt to sleep | Weekend | Past month |
| Bedtime | National Social Life, Health, and Aging Project (NSHAP) | What time do you usually go to bed and start trying to fall asleep? On weekdays or work days? [HH:MM am/pm] | Attempt to sleep | Workday or weekday |  |
| Bedtime | National Social Life, Health, and Aging Project (NSHAP) | What time do you usually go to bed and start trying to fall asleep? On weekends or days off? [HH:MM am/pm] | Attempt to sleep | Free |  |
| Bedtime | Health and Aging in Africa | Over the past 4 weeks, what time did you usually turn the lights off to go to sleep? [HH:MM am/pm] | Attempt to sleep |  | Past 4 weeks |
| Bedtime | Family Life, Activity, Sun, Health, and Eating (FLASHE) Study | What time do you usually go to bed in the evening (turn out the lights in order to go to sleep) on WEEKDAYS? [HH:MM am/pm] | Attempt to sleep | Weekday |  |
| Bedtime | Family Life, Activity, Sun, Health, and Eating (FLASHE) Study | What time do you usually go to bed in the evening (turn out the lights in order to go to sleep) on WEEKENDS? [HH:MM am/pm] | Attempt to sleep | Weekend |  |
| Bedtime | Sleep Heart Health Study (SHHS) | At what time do you fall asleep? [HH:MM am/pm] | Fall asleep time |  |  |
| Bedtime | uMCTQ | On WORKDAYS I normally fall asleep at __:__ AM/PM (this is NOT when you get into bed!) | Fall asleep time | Workday |  |
| Bedtime | uMCTQ | On WORK-FREE DAYS I normally fall asleep at __:__ AM/PM (this is NOT when you get into bed!) | Fall asleep time | Work-free |  |
| Bedtime | Heart Biomarker Evaluation in Apnea Treatment Study | At what time do you usually FALL ASLEEP on weekdays or your work days? [HH:MM am/pm] | Fall asleep time | Workday or weekday |  |

| Category | Study | Question Wording [Response Scale] | Event Definition | Context | Timeframe |
| --- | --- | --- | --- | --- | --- |
| Bedtime | Heart Biomarker Evaluation in Apnea Treatment Study | At what time do you usually FALL ASLEEP on weekends or your non-work days? [HH:MM am/pm] | Fall asleep time | Non-work day or weekend |  |
| Bedtime | Twin Cities Metropolitan Area 2000 Travel Behavior Inventory | What time did<THEY>go to sleep up on<THEIR>travel day? [HH:MM am/pm] | Fall asleep time |  | Travel day |
| Bedtime | The Common Cold Project | Time went to sleep last night? [HH:MM]am/pm | Fall asleep time |  | Last night |
| Bedtime | Korean General Social Survey (KGSS) | At about what time did you go to sleep yesterday? [HH:MM am/pm] | Fall asleep time |  | Yesterday |
| Bedtime | Biennial Media Consumption Survey | On a regular weeknight, what time do you usually go to sleep? [HH:MM am/pm] | Fall asleep time | Weeknight |  |
| Bedtime | Family Exchanges Study Wave 2, Philadelphia, Pennsylvania | What time did you go to sleep last night? [HH:MM am/pm] | Fall asleep time |  | Last night |
| Bedtime | Munich Chronotype Questionnaire (MCTQ) | On nights before workdays, I go to bed at _____ o clock ... | Time into bed | Workday |  |
| Bedtime | Munich Chronotype Questionnaire (MCTQ) | On nights before free days, I go to bed at _____ o clock ... | Time into bed | Free day |  |
| Bedtime | MrOS Sleep Study (MrOS) | During the past month, what time have you usually gone to bed at night? [HH:MM am/pm] | Time into bed |  | Past month |
| Bedtime | National Sleep Foundation: 2002 Adult Sleep Habits | At what time do you usually go to bed on nights before workdays or weekdays? [12:00am; 12:01-4:49am; 5:00am-5:29am; 5:30-5:59am; 6:00am-6:29am; 6:30-6:59am; 7:00-7:29am; 7:30-7:59am; 8:00-8:29am; 8:30-8:59am; 9:00-9:29am;9:30-9:59am;10:00-10:59am;11:00-11:59am;12:00-5:59pm;6-11:59pm] | Time into bed | Before workday or weekday |  |
| Bedtime | National Sleep Foundation: 2002 Adult Sleep Habits | At what time do you usually go to bed on nights you do not work the next day or weekends? [HH:MM am/pm] | Time into bed | Not work the next day or weekend |  |
| Bedtime | National Sleep Foundation: 2003 Sleep and Aging | At what time do you usually go to bed on weeknights? [HH:MM am/pm] | Time into bed | Weeknight |  |
| Bedtime | National Sleep Foundation: 2005 Adult Sleep Habits | At what time do you usually go to bed on nights you do not work the next day or weekends? [HH:MM am/pm] | Time into bed | Not work the next day or weekend |  |
| Bedtime | National Sleep Foundation: 2011 Sleep and Technology | What time do you usually go to bed? [HH:MM]am/pm | Time into bed |  |  |
| Bedtime | National Sleep Foundation: 2015 Sleep and Pain | Go to bed on work days or weekdays? [HH:MM am/pm] | Time into bed | Workday or weekday |  |
| Bedtime | National Sleep Foundation: 2015 Sleep and Pain | Go to bed on non-work days or weekends? [HH:MM am/pm] | Time into bed | Non-work day or weekend |  |
| Bedtime | Hispanic Community Health Study/Study of Latinos | The following two questions refer to the times you get in and out of bed in order to sleep (not including naps). What time do you usually go to bed? On weekdays? [HH:MM am/pm] | Time into bed | Weekday |  |
| Bedtime | Hispanic Community Health Study/Study of Latinos | The following two questions refer to the times you get in and out of bed in order to sleep (not including naps). What time do you usually go to bed? On weekends? [HH:MM am/pm] | Time into bed | Weekend |  |
| Bedtime | Apnea Positive Pressure Long-Term Efficacy Study (APPLES) | At what time do you usually FALL ASLEEP on weekends or your non-workdays? [ HH:MM am/pm] | Time into bed | Weekend |  |
| Bedtime | Apnea Positive Pressure Long-Term Efficacy Study (APPLES) | At what time do you usually FALL ASLEEP on weekdays or your workdays? [HH:MM am/pm] | Time into bed | Weeknight |  |
| Bedtime | Survey of Midlife in Japan (MIDJA 2) | During the past month, when have you usually gone to bed at night? [HH:MM am/pm] | Time into bed |  | Past month |
| Bedtime | Basic Nordic Sleep Questionnaire (BNSQ) | At what time do you usually go to bed (in order to sleep) during the working week? [HH:MM] | Time into bed | Working week |  |
| Bedtime | Basic Nordic Sleep Questionnaire (BNSQ) | At what time do you usually go to bed (in order to sleep) during free days? [HH:MM] | Time into bed | Free |  |
| Bedtime | Sleep Timing Questionnaire (STQ) | (Please think of GOOD NIGHT TIME as the time at which you are finally get in bed trying to fall asleep). On the night before a workday or school day, what is your earliest GOOD NIGHT TIME? [HH:MM am/pm] | Attempt to sleep | Work or school day |  |
| Bedtime | Sleep Timing Questionnaire (STQ) | (Please think of GOOD NIGHT TIME as the time at which you are finally get in bed trying to fall asleep). On the night before a workday or school day, what is your latest GOOD NIGHT TIME? [HH:MM am/pm] | Attempt to sleep | Work or school day |  |
| Bedtime | Sleep Timing Questionnaire (STQ) | (Please think of GOOD NIGHT TIME as the time at which you are finally get in bed trying to fall asleep). On the night before a workday or school day, what is your usual GOOD NIGHT TIME? [HH:MM am/pm] | Attempt to sleep | Work or school day |  |
| Bedtime | Sleep Timing Questionnaire (STQ) | (Please think of GOOD NIGHT TIME as the time at which you are finally get in bed trying to fall asleep). On the night before a day off (e.g., weekend), what is your earliest GOOD NIGHT TIME? [HH:MM am/pm] | Attempt to sleep | Day off or weekend |  |
| Bedtime | Sleep Timing Questionnaire (STQ) | (Please think of GOOD NIGHT TIME as the time at which you are finally get in bed trying to fall asleep). On the night before a day off (e.g., weekend), what is your latest GOOD NIGHT TIME? [HH:MM am/pm] | Attempt to sleep | Day off or weekend |  |
| Bedtime | Sleep Timing Questionnaire (STQ) | (Please think of GOOD NIGHT TIME as the time at which you are finally get in bed trying to fall asleep). On the night before a day off (e.g., weekend), what is your usual GOOD NIGHT TIME? [HH:MM am/pm] | Attempt to sleep | Day off or weekend |  |
| Wake time | MrOS Sleep Study (MrOS) | During the past month, when have you usually gotten up in the morning?" [HH:MM am/pm] | Get up |  | Past month |
| Wake time | National Sleep Foundation: 2005 Adult Sleep Habits and Styles | At what time do you usually get up on days you do not work or weekends?" [HH:MM am/pm] | Get up | Non-work day or weekend |  |
| Wake time | Munich Chronotype Questionnaire (MCTQ) | On workdays... I have to get up at _____ o'clock | Get up | Work |  |
| Wake time | Survey of Midlife in Japan (MIDJA 2) | During the past month, when have you usually gotten up in the morning? [HH:MM am/pm] | Get up |  | Past month |
| Wake time | National Sleep Foundation: 2002 Adult Sleep Habits | Thinking about your usual non-workday or weekend, please answer the following questions. At what time do you usually get up on days you do not work or weekends? [12:00am; 12:01-4:49am; 5:00am-5:29am; 5:30-5:59am; 6:00am-6:29am; 6:30-6:59am; 7:00-7:29am; 7:30-7:59am; 8:00-8:29am; 8:30-8:59am; 9:00-9:29am; 9:30-9:59am; 10:00-10:59am; 11:00-11:59am; 12:00-5:59pm; 6-11:59pm] | Get up | Before workday or weekday | Past 4 weeks |
| Wake time | National Sleep Foundation: 2002 Adult Sleep Habits | At what time do you usually get up on days you work or on weekdays? [12:00am; 12:01-4:49am; 5:00am-5:29am; 5:30-5:59am; 6:00am-6:29am; 6:30-6:59am; 7:00-7:29am; 7:30-7:59am; 8:00-8:29am; 8:30-8:59am; 9:00-9:29am; 9:30-9:59am; 10:00-10:59am; 11:00-11:59am; 12:00-5:59pm; 6-11:59pm] | Get up | Not work the next day or weekend |  |
| Wake time | Health and Aging in Africa | Over the past 4 weeks, what time did you usually get out of bed? [HH:MM am/pm] | Time out of bed |  |  |
| Wake time | Family Life, Activity, Sun, Health, and Eating (FLASHE) | What time do you usually get out of bed in the morning on WEEKDAYS? [HH:MM am/pm] | Time out of bed | Weekday |  |
| Wake time | Family Life, Activity, Sun, Health, and Eating (FLASHE) | What time do you usually get out of bed in the morning on WEEKENDS? [HH:MM am/pm] | Time out of bed | Weekend |  |
| Wake time | Sleep Heart Health Study (SHHS) | At what time do you wake up? [HH:MM am/pm] | Wake time |  |  |
| Wake time | National Sleep Foundation: 2011 Sleep and Technology | What time do you wake up? [HH:MM am/pm] | Wake time |  |  |
| Wake time | National Sleep Foundation: 2015 Sleep and Pain | Wake up on work days or weekdays? [HH:MM am/pm] | Wake time | Workday or weekday |  |
| Wake time | National Sleep Foundation: 2015 Sleep and Pain | Wake up on non-work days or weekends? [HH:MM am/pm] | Wake time | Non-work day or weekend |  |
| Wake time | Hispanic Community Health Study/Study of Latinos | The following two questions refer to the times you get in and out of bed in order to sleep (not including naps). What time do you usually wake up? On weekends? [HH:MM am/pm] | Wake time | Weekend |  |
| Wake time | Hispanic Community Health Study/Study of Latinos | The following two questions refer to the times you get in and out of bed in order to sleep (not including naps). What time do you usually wake up? On weekdays? [HH:MM am/pm] | Wake time | Weekday |  |
| Wake time | HeartBEAT (Heart Biomarker Evaluation in Apnea Treatment) Study | At what time do you usually WAKE UP on weekdays or your work days? [HH:MM am/pm] | Wake time | Weekday |  |
| Wake time | HeartBEAT (Heart Biomarker Evaluation in Apnea Treatment) Study | At what time do you usually WAKE UP on weekends or your non-work days?" [HH:MM am/pm] | Wake time | Non-work day or weekend |  |
| Wake time | Apnea Positive Pressure Long-Term Efficacy Study (APPLES) | At what time do you usually WAKE UP on weekends or your non-work days? [HH:MM am/pm] | Wake time | Weekend |  |
| Wake time | Apnea Positive Pressure Long-Term Efficacy Study (APPLES) | At what time do you usually WAKE UP on weekdays or your work days? [HH:MM am/pm] | Wake time | Weeknight |  |
| Wake time | Cleveland Family Study | What time do you did you wake up from your usual sleep on weekdays? [HH:MM am/pm] | Wake time | Weekday |  |
| Wake time | Cleveland Family Study | What time do you did you wake up from your usual sleep on weekends? [HH:MM am/pm] | Wake time | Weekend |  |
| Wake time | Munich Chronotype Questionnaire (MCTQ) | On free days (please only judge normal free days, ie, without parties etc) I normally wake up at _____ o clock | Wake time | Free |  |
| Wake time | uMCTQ | On WORKDAYS I normally wake up at __:__ AM/PM (this is NOT when you get out of bed!) | Wake time | Workday |  |
| Wake time | uMCTQ | On WORK-FREE DAYS when I don't use an alarm I normally wake up at __:__ AM/PM (this is NOT when you get out of bed!) | Wake time | Work-Free |  |
| Wake time | Twin Cities Metropolitan Area 2000 Travel Behavior Inventory | Wake onset: What time did<THIEY>wake up on<THIER>travel day? [HH:MM am/pm] | Wake time |  |  |
| Wake time | National Social Life, Health, and Aging Project (NSHAP) | What time do you usually wake up? On weekdays or work days? [HH:MM am/pm] | Wake time | Workday or weekday |  |
| Wake time | National Social Life, Health, and Aging Project (NSHAP): | What time do you usually wake up? On weekends, or days off? [HH:MM am/pm] | Wake time | Days off or weekend |  |
| Wake time | CBS News Monthly Poll | What time do you usually wake up on a typical Sunday morning? [before 6am; between 6am and 659am; between 7am and 759am; between 8 and 859 am; between 9 and 959am; between 10 and 1059am; 11am or later; Work nights so sleep all day] | Wake time | Sunday morning |  |
| Wake time | Basic Nordic Sleep Questionnaire (BNSQ) | At what time do you usually wake up during the working week? [HH:MM] | Wake time | Working week |  |
| Wake time | Basic Nordic Sleep Questionnaire (BNSQ) | At what time do you usually wake up during free days? [HH:MM] | Wake time | Free |  |
| Wake time | Sleep Timing Questionnaire (STQ) | (Please think of GOOD MORNING TIME as the time at which you finally get out of bed and start your day). Before a work day or school day, what is your earliest GOOD MORNING TIME? [HH:MM am/pm] | Wake time | Work or school day |  |
| Wake time | Sleep Timing Questionnaire (STQ) | (Please think of GOOD MORNING TIME as the time at which you finally get out of bed and start your day). Before a work day or school day, what is your latest GOOD MORNING TIME? [HH:MM am/pm] | Wake time | Work or school day |  |
| Wake time | Sleep Timing Questionnaire (STQ) | (Please think of GOOD MORNING TIME as the time at which you finally get out of bed and start your day). Before a work day or school day, what is your usual GOOD MORNING TIME? [HH:MM am/pm] | Wake time | Work or school day |  |
| Wake time | Sleep Timing Questionnaire (STQ) | (Please think of GOOD MORNING TIME as the time at which you finally get out of bed and start your day). Before a day of (e.g., a weekend), what is your earliest GOOD MORNING TIME? [HH:MM am/pm] | Wake time | Free or weekend day |  |
| Wake time | Sleep Timing Questionnaire (STQ) | (Please think of GOOD MORNING TIME as the time at which you finally get out of bed and start your day). Before a day of (e.g., a weekend), what is your latest GOOD MORNING TIME? [HH:MM am/pm] | Wake time | Free or weekend day |  |
| Wake time | Sleep Timing Questionnaire (STQ) | (Please think of GOOD MORNING TIME as the time at which you finally get out of bed and start your day). Before a day of (e.g., a weekend), what is your usual GOOD MORNING TIME? [HH:MM am/pm] | Wake time | Free or weekend day |  |

*Notes:*
A blank cell indicates that this component is not present in the question.
The sentence case (e.g., all capitals) is displayed as represented on the survey.

We identified 36 distinct questions assessing wake times. Regarding event definitions, 27 questions asked participants to report the time they wake up (e.g., Basic Nordic Sleep Questionnaire: “At what time do you usually wake up during the working week?”),^21^ while 6 surveys assessing wake times asked participants to report the time they “get up” (e.g., National Sleep Foundation – 2005 Adult Sleep Habits and Styles: “At what time do you usually get up on days you do not work or weekends?”)^22^ and 3 questions asked participants to report their out of bed time (FLASHE Study: “What time do you usually get out of bed in the morning on weekdays”).Thirty of the wake time questions asked participants to distinguish between work- and free- days (e.g.: Heart Biomarker Evaluation in Apnea Treatment study: “At what time do you usually WAKE UP on weekends or your non-work days?”). Three questions provided participants with a timeframe within which to report their wake times (e.g., Health and Aging in Africa: A Longitudinal Study of an In-Depth Community in South Africa: “Over the past 4 weeks, what time did you usually get out of bed?”).^24^

We found one question that asked participants to report their method of waking (the uMCTQ:“On work-free days when I don’t use an alarm clock I normally fall asleep at ___”).^16^

### Sleep Latency (Table 4)

We identified 15 distinct questions assessing sleep latency. The event definition for latency was uniform across questions (e.g., “minutes to fall asleep”). Five sleep latency questions asked participants to distinguish between work- and free-day sleep latency (e.g., Munich Chronotype Questionnaire: “On nights before workdays, it takes me ___mins to fall asleep”) and 3 asked participants to report their sleep latency in a specific timeframe (e.g., Sleep Heart Health Study: “During the past month, how long (in minutes) has it usually taken you to fall asleep each night?”).^25^

**Table 4.** Sleep latency questions.

| **Category** | **Study** | **Question Wording [Response Scale]** | **Context** | **Timeframe** |
| --- | --- | --- | --- | --- |
| Sleep Latency | Sleep Heart Health Study (SHHS) | During the past month, how long (in minutes) has it usually taken you to fall asleep each night? [MM] |  | Past month |
| Sleep Latency | National Sleep Foundation: 2005 Adult Sleep Habits and Styles | How long, on most nights, does it take you to fall asleep? [HH:MM] |  |  |
| Sleep Latency | Framingham Heart Study | How long does it usually take you to fall asleep at bedtime? [HH:MM] |  |  |
| Sleep Latency | Survey of Midlife in Japan (MIDJA 2) | During the past month, how long (in minutes) has it taken you to fall asleep at night? |  | Past month |
| Sleep Latency | National Health and Nutrition Examination Survey (NHANES) | How long does it usually take to fall asleep at bedtime? |  |  |
| Sleep Latency | Munich Chronotype Questionnaire (MCTQ) | On nights before workdays it takes me _____ mins to fall asleep | Work |  |
| Sleep Latency | Munich Chronotype Questionnaire (MCTQ) | On nights before free days it takes me _____ mins to fall asleep | Free |  |
| Sleep Latency | Munich Chronotype Questionnaire (MCTQ) | On nights before free days I generally fall asleep after no more than _____ min | Free |  |
| Sleep Latency | Pittsburgh Sleep Quality Index (PSQI) | How long (in minutes) has it taken you to fall asleep each night? |  |  |
| Sleep Latency | Sleep Timing Questionnaire (STQ) | On most nights, how long, on average does it take you to fall asleep after you start trying? _____ minutes |  |  |
| Sleep Latency | St Mary's Hospital Sleep Questionnaire | How long did it take you to fall asleep last night? [HH:MM] |  | Last night |
| Sleep Latency | Medical outcomes Study Sleep Scale (MOS-SS) | How long did it take you to fall asleep during the past 4 weeks? [1: 0-15minutes; 2: 16-30minutes; 3: 31-45 minutes; 4: 46-60 minutes; 5: More than 60 minutes] |  | Past 4 weeks |
| Sleep Latency | Basic Nordic Sleep Questionnaire (BNSQ) | How long time (how many minutes as an average) do you stay awake in bed before you fall asleep (after lights off)? During the working days it takes me about [MM] minutes before I fall asleep. | Work |  |
| Sleep Latency | Basic Nordic Sleep Questionnaire (BNSQ) | How long time (how many minutes as an average) do you stay awake in bed before you fall asleep (after lights off)? During free time it takes me about [MM] minutes before I fall asleep. | Free |  |
| Sleep Latency | Multi-Ethnic Study of Atherosclerosis (MESA) | How long does it usually take you to fall asleep at bedtime? |  |  |

*Notes:*
A blank cell indicates that this component is not present in the question.
The sentence case (e.g., all capitals) is displayed as represented on the survey.
Sleep latency questions were all similarly defined, therefore there is no “Event Definition” column.

### Napping (Table 5)

We identified 8 distinct napping questions. Among the napping questions, three asked participants to report the number of naps that were “5 minutes or longer” while 1 question asked participants to report the number of naps that were “an hour or two.” Regarding context, one question asked participants to report work-versus free-day napping (e.g., Sleep Disorders Questionnaire: “How many daytime naps (asleep for 5 minutes or more) do you take on an average working day?”);^13^ Four provided participants with a timeframe, such as the past week (e.g., National Social Life, Health, and Aging Project: “During the past week, on how many days did you nap for 5 minutes or more”).^26^

**Table 5.** Napping questions.

| **Category** | **Study** | **Question Wording [Response Scale]** | **Event Definition** | **Context** | **Timeframe** |
| --- | --- | --- | --- | --- | --- |
| Naps | Hispanic Community Health Study/Study of Latinos | During a usual week, how many times do you nap for 5 minutes or more? [none; 1 or 2 times; 3 or 4 times; 5 or more times] | Nap frequency (defined as 5 mins) |  | Usual week |
| Naps | National Social Life, Health, and Aging Project (NSHAP) | During the past week, on how many days did you nap for 5 minutes or more? [never; 1 or 2 days; 3 or 4 days; 5 or more days] | Nap frequency (defined as 5 mins) |  | Past week |
| Naps | National Social Life, Health, and Aging Project (NSHAP) | During the past week, on how many days did you nap for an hour or two? [never; 1 or 2 days; 3 or 4 days; 5 or more days] | Nap frequency (defined as 1-2 hours) |  | Past week |
| Naps | Sleep Disorders Questionnaire | How many daytime naps (asleep for 5 minutes or more) do you take on an average working day? [1.) None 2.) One 3.) Two 4.) Three or four 5.) Five or more] | Nap frequency minutes (defined as 5 minutes or longer) | Working day |  |
| Naps | Basic Nordic Sleep Questionnaire (BNSQ) | How often do you sleep naps at daytime? [1: never or less than once per week; 2: less than once per week; 3: on 1-2 days per week; 4: on 3-5 days per week; 5: daily or almost daily] | Frequency of any nap |  | Per week |
| Naps | Basic Nordic Sleep Questionnaire (BNSQ) | If you sleep a nap, how long does it last for? My naps usually last for about [HH:MM] | Duration of nap |  |  |
| Naps | Multi-Ethnic Study of Atherosclerosis (MESA) | How often do you "make time" in your schedule for a regular nap or "siesta" in the afternoon? [Never or rarely; Sometimes; Often; Every day or almost every day] | Frequency of any nap |  |  |
| Naps | Multi-Ethnic Study of Atherosclerosis (MESA) | When you do nap in the afternoon, how long do you sleep? [HH:MM]\ | Duration of nap |  |  |

*Notes:*
A blank cell indicates that this component is not present in the question.
The sentence case (e.g., all capitals) is displayed as represented on the survey.

### Critiquing approaches (Table 6)

In Table 6, we address the advantages and disadvantages of different event definition(s), context, and timeframe considerations of questions within each of the five domains to identify word choices that influence the user’s response. The most important are distinguishing between different event definitions of bed and wake times (e.g., when a person “gets up” versus “wakes up”) and of overall sleep timing (i.e., whether nighttime sleep is assumed to be the only sleep episode); whether or not a timeframe (e.g., “in the past 4 weeks”) is provided; and segregating responses for work-and free-days.

**Table 6.** Sleep duration and timing question design considerations.

Sleep duration and timing question design considerations.
| Component of Interest | Example(s) | Design Considerations |
| --- | --- | --- |
| Event definition<br>(sleep duration) | "How many hours do you **usually sleep at night**?"<br>"How much sleep do you usually get at night (or your **main sleep episode**)?"<br>**Time in bed attempting to sleep** (calculated from questions assessing the first "attempt to sleep" and the time "out of bed after waking")<br>**Major sleep episode** (calculated from questions assessing the first "attempt to sleep," the "time it takes to fall asleep," and "the time a person wakes up") | Reporting nighttime sleep may be challenging for shift workers who commonly sleep during the day. Requesting information about "main sleep episode" (as opposed to "at night") would enable shift workers and others on non-traditional schedules to respond with their primary sleep episode.<br>Instead of asking for sleep duration directly, researchers can also ask for sleep times in order to calculate sleep duration. Researchers interested in the time spent in bed close to sleep (herein proposed as "<u>time in bed attempting to sleep</u>") may administer questions assessing the first attempt to sleep and the time out of bed in the morning. On the other hand, using the time a person reports "going to bed" and "getting out of bed" (common questions administered to measure time in bed) may be extremely protracted for individuals who spend extended periods of time in bed before sleep. Researchers interested in measuring the <u>major sleep episode</u> may administer questions assessing the times a person "first attempts to sleep," how long it takes to "fall asleep," and when the person "wakes up." |
| Context<br>(all domains) | "How much sleep do you get at night on **weekdays or workdays**?"<br>"How much sleep do you get at night on **weekends or free days**?" | Requesting information about week versus work days allows participants to respond specifically if the sleep timing or duration differs between the two conditions.<br>Asking participants to distinguish between sleep on week/weekend nights may not capture accurate timing details for people who work on weekends. |
| Time Frame<br>(all domains) | "On average how many hours did you sleep each night during **the past 4 weeks**?"<br>"**During the past month**, what time have you usually gone to bed at night?" | Asking participants to report their "usual" sleep duration and/or timing may affect the quality of responses because individuals are limited in their ability to report their behavior in an abstract time frame.<br>Providing a timeframe (e.g., "During the past month" or "In the past 2 weeks") guides participant responses. |
| Response option<br>(all domains) | "**HH:MM**"<br>"[<5 hours; hrs; 6 hrs; 7 hrs; 8 hrs; 9 hrs; 10+ hours]" | The HH:MM response option enables more precise recording of actual duration.<br>Individuals may differ in how precise their responses are (e.g., every one minute or rounding to quarter hour).<br>Closed-ended scales may be beneficial for reducing the survey completion time, |
| Event definition<br>(bedtime) | "At what time do you **first attempt to sleep**?"<br>At what time do you **fall asleep**?"<br>At what time do you **go to bed**?" | "First attempt to sleep" is beneficial in that it asks precisely when the person first attempts sleep, which, in conjunction with sleep latency may be used to calculate sleep duration.<br>"Fall asleep" offers more precise guidance to the participant to report the time they fall asleep.<br>"Go to bed" is ambiguous; it is unclear whether the person should respond with the time |
|  |  | they get in bed or first attempt sleep. |
| Event definition<br>(wake time) | "At what time do you usually **get out of bed?**"<br>"At what time do you typically **get up?**"<br>"At what time do you usually **wake up?**" | "Get out of bed" is useful in capturing time in bed.<br>"Gets up" is ambiguous; this could mean when a person gets out of bed or wakes up.<br>"Wake up" is clear and assesses the time a person wakes from sleep. |
| Event definition | "During the past week, on how many days did you **nap for an hour or two?**"<br>"During the past week, on how many days did you nap for **5 minutes or more?**" | A question that asks for participants to report napping for one or two hours may capture longer naps but not shorter naps.<br>A question asking for naps "5 minutes or longer" will capture both short naps as well as long naps (without distinguishing between the two). |

*Note.*
Bold indicates the wording approaches a survey designer may consider.

## Discussion

Our methodological review includes 49 surveys that assess sleep duration and/or timing via self-reported survey (nine validated surveys and 40 non-validated surveys). In our analysis, we distinguish between sleep duration and timing-related domains, including questions assessing: sleep duration; bedtimes; wake times; sleep latency; and napping.

Our results uncover several primary findings regarding the common approaches to measuring sleep duration and/or timing via survey. First, we document significant semantic variation in the event definitions in each question. Across questionnaires, sleep duration is estimated using a variety of approaches, including single item questions regarding estimated total time asleep (e.g., during the night or across the 24-hour day) or potentially derived from differences in times reported for bedtimes or sleep onset times and wake times. Significant biases in sleep duration estimates obtained from questionnaire as compared to actigraphy have been reported, with the directionality (over or under-estimation) and magnitude of such biases varying according to the specific questions used.^27^

Sleep timing questions, including wake bed times and waketimes also varied widely between questionnaires. Questions most commonly asked participants to report the time they “get into bed,” while intuitively straight forward, can result in over-estimation of sleep duration, especially if the participant spends time in bed reading or watching television before sleeping. While the limitation of asking a participant the time they “get into bed” may be addressed by also asking about estimated sleep latency, both healthy sleepers and those with insomnia, vary widely in their ability to accurately report sleep latency, and those with perceived problems with sleep quality may particularly over-estimate sleep latency, resulting in differential bias.^28,29^ Asking about time “going to sleep” similarly may be biased by information recall bias, especially among individuals with perceived poor sleep quality.

We also detect interesting differences in assessment of wake times, including the time a person “wakes up” versus “gets out of bed.” Further, some questions simply asked when a participant typically “gets up,” which could be interpreted as either wake time or time out of bed. These small semantic nuances have important implications for the responses a participant is to provide. Asking a participant when they “get out of bed” is likely to overestimate sleep duration, particularly if a participant spends an extended amount of time in bed after waking but before getting out of bed (e.g., to check their emails), and may be appropriate for scoring time in bed. On the other hand, a question that asks a participant when they “wake up” may be more appropriate for researchers interested in capturing sleep duration as opposed to time in bed.

We were surprised to see only one question address the method of awakening. Information about whether the awakening is spontaneous – and therefore sleep ended for internal physiological reasons, rather than externally imposed (e.g., an alarm clock) – may be important for some outcomes. It is also not known how a person who uses the “snooze bar “ on an alarm clock (i.e., to go back to sleep and have the alarm sound later) may respond to a question that asks the time they “woke up:” such a person could respond with their first wake time or the last time they awoke after hitting the snooze bar once or multiple times.

Second, we document significant variation in context and timeframe cues. Despite the prevalence with which sleep schedules vary between work- and free-days (contributing to ‘social jetlag’^7^ and its adverse cardiometabolic consequence^30–33^) these context cues were frequently not presented in the questions. Another distinction is the way different days of the week were described. In some surveys, participants were asked to report their sleep on a “weekday” versus “weekend,” which may not yield expected results for individuals who work on weekends. Furthermore, for some people non-work days may not be entirely “free” days that allow more choice in sleeping timing and duration because of family, social, or other obligations. Therefore, researchers interested in sleep on a true “free” day may choose to ask participants to report sleep duration and/or timing on days they wake without an alarm or other external stimulus or obligation. Distinguishing between work and free day sleep allows for estimation of social jetlag, which increases risk of adverse outcomes^7,30–32,34^. Night-to-night variability of sleep timing, which is also a risk factor for adverse outcomes,^1,35^ cannot be calculated from these surveys.

Very few questions asked participants to report on their sleep in a specific timeframe, such as “In the past week, how many hours did you spend sleeping?” Instead, most questions asked how much individuals “usually sleep.” These language variants are important because behavioral research states that individuals are handicapped in their ability to report on an abstract time period, and the quality of question responses increases dramatically when participants are provided with a specific time period, such as the prior week or month.^36^ This is in accordance with recency bias, which refers to the fact that recall accuracy wanes as time increases between the time at which an event transpired and the time an individual is asked to recall the event.^37^ In comparison, the nutrition literature has critiqued even 24-hour recall of dietary intake as susceptible to reporting biases, arguing in favor of immediate recall using a diary method.^38^

Few surveys asked about naps and the definition of a nap varied widely between surveys. One survey asked participants to report the number of naps lasting five minutes or longer. Another survey asked specifically for the frequency of naps that lasted “an hour or two.” Although five-minute naps were used frequently in questions, the authors cannot identify literature that affirms the appropriateness of using a five-minute definition for a nap. Measuring longer nap periods may be useful for a researcher interested in measuring sleep across the 24-hour day, while shorter naps may be of interest to researchers studying markers of excessive daytime sleepiness, for which short naps are an indicator. Another approach was to ask participants to simply report how many minutes they typically spend napping.^21^ Among the surveys that did ask about naps, only some included a timeframe (e.g., “in the past 4 weeks"). Variation in the duration of naps may represent both differences in physiological needs and behaviors, as well as result in different physiological responses (e.g., extent to which deep sleep is achieved). Planned napping (e.g., “siestas” or scheduled naps as “counter-measures” before night work) as compared to unintentional napping also may have profoundly different health implications. Yet, these differences were not captured in the surveys reviewed in this report.

Most studies identified in this review measured nocturnal sleep, which is not appropriate for shift workers who typically sleep during the daytime and represent approximately 15% of the population.^39–41^ For instance, approximately half of the sleep duration questions asked participants to report their sleep “at night.” A potential alternate approach was to ask participants to report their “longest sleep episode,” although this wording may be difficult for those who are not shift workers to comprehend.

Our study is the first to provide a rigorous methodological review of common approaches to measuring self-reported sleep duration and/or timing. We here demonstrate the lack of consensus on event definition(s), context, and timeframe-related considerations in self-reported sleep duration and/or timing questions.

Our research offers question guidance and design considerations for researchers and developers of new survey instruments to assess sleep duration and/or timing. First, the choice of survey questions must be guided by the research objective. For instance, a researcher interested in measuring sleep duration may choose to phrase bedtime and wake time questions to assess actual sleep rather than assess time in bed though time into bed and time out of bed questions t. This would recognize the potential differential and non-differential biases that may result from use of alternative wording (e.g., related to sleep vs. time in bed). Furthermore, “time in bed” is itself potentially ambiguous, particularly important with recent trends toward working from home as well as COVID-19 stay-at-home recommendations that increase the opportunity for extended periods of time in their bed even when not attempting to sleep. More specific wording may be “time in bed attempting to sleep.” This was used in 2016 Consumer Technology Association report for establishing standards on sleep wearable devices^42^ that defines bedtime to be the “time when an individual began attempting to sleep.” Second, there may be limits on the number of questions available in the survey; under such conditions, researchers may consider asking directly for sleep duration as opposed to the time points that would enable its calculation. Finally, researchers and industry representatives may work together for comparing self-reported sleep measurements with commercial sleep-related products and services.

Studies have reported that responses to sleep questions (compared to actigraphy) may differ across the population, with variation by ethnicity/race, presence/absence of insomnia, education level and other factors.^43^ Additional methodological research is needed to ensure that the choice of specific questions is most appropriate for the population under study, and appropriate instructions and contextual information are provided to minimize misinterpretation and bias.

This work has several limitations. We only reviewed English language questions pertaining to adults and we did not have data on the total number of surveys using each question type.

An area for future research is to consider the implications for different populations of the different approaches to measuring sleep duration and timing. The relationship to different wording approaches described above and objective sleep parameters varies significantly between ethnic groups.^27,43^ Future research should examine language, cultural or health literacy factors relating to sleep duration and timing questions. Future research may also consider differences in question event definition(s), context, and timeframe during the validation procedure.

## Conclusion

Our review highlights the large variation in approaches for ascertaining sleep duration and timing across published sleep questionnaires. We document several significant differences in measuring sleep duration and timing and their related domains, including sleep duration, bedtimes, wake times and/or method of awakening, sleep latency, and napping. As public health recommendations are often derived from data from surveys employing different survey approaches, future work to promote consensus methodology for measurement of sleep duration and timing may be beneficial. A major challenge is to develop a framework for validating sleep questionnaires which assesses the sleep-related domains are most relevant for public health or clinical use. Finally, it will be critical that questionnaire development and validation considers their use across populations that vary by demographic factors, work histories, geography and clinical conditions.

## Data Availability

Data is available upon request.

## Conflicts of Interest

RR reports personal fees from Denihan Hospitality, SleepCycle, Rituals Cosmetics, and byNacht. SFQ reports research funding from the National Institutes of Health, serves as a consultant to Jazz Pharmaceuticals, Whispersom and is a committee chair and a taskforce member for the American Academy of Sleep Medicine. SR receives grants or contracts funding from the NIH, Department of Defense, and Jazz Pharmaceuticals and has consulted for Jazz Pharmaceuticals, Respicardia Inc, and Eisai Pharmaceuticals and serves on committees for the American Thoracic Society, National Sleep Foundation, and American Academy of Sleep Medicine. MDW reports research support from NIOSH and the Brigham Research Institute, and has consulted for the University of Pittsburgh and the National Sleep Foundation. E.B.K. has received travel reimbursement from the Sleep Research Society, the National Sleep Foundation, the Santa Fe Institute, the World Conference of Chronobiology, and the Gordon Research Conference; she was paid by the Puerto Rico Trust for a grant review, and has consulted for the National Sleep Foundation. CAC is/was a paid consultant to Bose, Boston Celtics, Boston Red Sox, Cephalon, Institute of Digital Media and Child Development, Klarman Family Foundation, Jazz Pharma, Merck, Purdue Pharma, Samsung, Teva Pharma Australia, AARP, American Academy of Dental Sleep Medicine, Eisenhower Medical Center, M. Davis and Company, Physician’s Seal, UC San Diego, University of Washington, University of Michigan, Maryland Sleep Society, National Sleep Foundation, Sleep Research Society, Tencent, and Vanda Pharmaceuticals, in which Dr. Czeisler also holds an equity interest; receives research/education support through BWH from Cephalon, Mary Ann & Stanley Snider via Combined Jewish Philanthropies, NFL Charities, Jazz Pharmaceuticals Plc Inc, Philips Respironics Inc, Regeneron Pharmaceuticals, Teva Pharmaceuticals Industries Ltd, Sanofi SA, Optum, ResMed, San Francisco Bar Pilots, Sanofi, Schneider, Simmons, Sysco, Philips, Vanda Pharmaceuticals; is/was an expert witness in legal cases, including those involving Advanced Power Technologies, Alvarado Hospital, LLC, Amtrak; Bombardier, Inc., C&J Energy Services; Casper Sleep, Inc., Columbia River Bar Pilots, Complete General Construction Company, Dallas Police Department, Delta Airlines/Comair, Enterprise Rent-A-Car, Fédération des Medecins Residents du Quebec (FMRQ), FedEx, Greyhound Lines, Inc./Motor Coach Industries/FirstGroup America, H.G. Energy LLC, Maricopa County, Arizona, Sheriff’s Office, Murrieta Valley Unified School District, Pomerado Hospital, Palomar Health District, Puckett EMS, Purdue Pharma, South Carolina Central Railroad Company, LLC., Steel Warehouse, Inc., Union Pacific Railroad, United Parcel Service, and Vanda Pharmaceuticals; serves as the incumbent of an endowed professorship provided to Harvard University by Cephalon, Inc. CAC receives royalties from McGraw Hill, and Philips Respironics for the Actiwatch-2 and Actiwatch Spectrum devices. Dr. Czeisler’s interests were reviewed and are managed by Brigham and Women’s Hospital and Partners HealthCare in accordance with their conflict of interest policies.

## Acknowledgements

NIH NHLBI National Sleep Research Resource, contract# K01HL150339 (RR), 75N92019C00011 (SR, YZ); K24-HL105664 (EBK); P01-AG009975 (CAC, EBK), R01-HL128538 (EBK), U54-AG062322 (EBK). NIOSH R01 OH011773 (LKB, MDW). Brigham Research Institute Fund to Sustain Research Excellence (MDW).

## Notes

### Author Declarations

This study did not include human subjects.

